# Cortical surface area drives volumetric and cognitive deficits in complex congenital heart disease

**DOI:** 10.1101/2025.10.09.25337644

**Authors:** Asuka Toyofuku, Melanie Ehrler, Oliver Kretschmar, Beatrice Latal, Ruth O’Gorman Tuura

## Abstract

Individuals with complex congenital heart disease (CHD) are at increased risk for cognitive impairments linked to altered brain structure, including reduced cortical volume. Cortical volume is determined by two distinct morphometric measures: cortical thickness (CT) and surface area (SA), each with unique developmental, genetic, and evolutionary origins. Despite their importance, few studies have examined the differential contributions of CT and SA to cortical volume alterations and cognitive outcomes in CHD. Exploring these individual cortical morphometric changes may provide additional insight into the origin of cortical volume changes in CHD, and their relationships with cognitive function.

High-resolution 3D T1-weighted images, IQ and executive function (EF) scores were acquired from a final sample of 49 patients with CHD and 80 controls (mean age=13.16, male:54.3%). Cortical reconstruction and volumetric analyses were performed using Freesurfer version 7.1. Group differences in cortical volume, CT, and SA, as well as their associations with cognitive measures, were investigated through vertex-wise multivariate general linear modelling.

While both SA and CT were significantly reduced in the CHD group, both globally and regionally, SA was more affected than CT, and total SA showed a stronger and more consistent correlation with cortical volume. Furthermore, SA, but not CT, was strongly associated with IQ and EF across the brain. These relationships were especially prominent in the frontal and occipitotemporal cortices, where patients with CHD exhibited stronger associations between SA and cognitive performance compared to controls.

These findings indicate that cortical volume reductions observed in CHD primarily reflect reduced cortical surface area rather than cortical thinning. While structure-function associations between cortical morphology and cognition are established in healthy populations, our data suggest that these relationships are accentuated in CHD, likely due to disrupted neurodevelopmental processes. This study underscores the importance of differentiating cortical morphometric features to improve our understanding of brain-behaviour associations and the neurobiological underpinnings of cognitive impairment in CHD.ss

## Introduction

Congenital heart disease (CHD) refers to a range of structural abnormalities of the heart or its major blood vessels, affecting approximately 8 per 1,000 live births.^1^ Due to altered brain perfusion, chronic hypoxia, brain injuries and many other clinical risk factors, individuals with CHD are often reported to have a spectrum of cognitive impairments together with altered brain structures — ranging from total and regional grey/white matter volume reduction, white matter microstructure integrity and structural connectivity.^2,3^ Structural imaging studies have suggested that populations with CHD already show smaller brain volumes during the fetal period,^4–9^ throughout the neonatal and postnatal periods,^10–17^ and this gap persists until adulthood.^18–21^ The majority of these studies showed that smaller brain volumes were attributable to smaller cortical volumes among other regional volume reductions.

However, the cortical volume, which the majority of CHD brain-imaging studies assessed, is computed via multiplication of cortical thickness (CT) and surface area (SA). CT is defined as the distance between the pial surface and the white matter surface, while SA is the area of the pial surface covering all gyri and sulci. CT and SA are suggested to be evolutionarily distinct^22,23^ and controlled by different genetic variants,^24,25^ highlighting their independent developmental trajectories. Given that volumetric findings could reflect any combination of altered CT or altered SA,^26^ investigating CT, SA, and volume together can lend additional insight into individual differences in brain structure^27^ and may shed light on the mechanistic underpinnings of brain alterations in patients with CHD.

So far, morphometric studies on patients with CHD are relatively scarce, but a few studies have been conducted in fetuses,^5,28^ neonates^16,29^ and adolescents,^30^ which all reported a reduction of total SA without regional predilections, except for one fetal study that reported no CHD-control difference.^28^ One adolescent study investigated SA in a lobe-based manner, reporting statistically significant SA reductions for the CHD group in temporal and parietal lobes, less so in frontal lobes, but not in occipital lobes.^20^ In contrast to the relatively consistent report on reduced total SA, findings on mean CT in CHD are conflicting. One study in neonates^16^ and another in adolescents^20^ found no statistically significant group difference in mean CT, while another adolescent/adult study found significantly decreased mean CT.^30^ Another study examined regional CT in 10 adults with CHD and reported a widespread bilateral decrease in CT.^31^

To date, whole brain analyses of regional SA and CT changes in children/adolescents with CHD have not been reported in the literature, and it is not yet known whether volume reductions are predominantly due to reduced SA or CT, or both. Furthermore, the cortical structure-function relationship and the distinct role played by CT and SA in cognitive function in CHD are under-researched. Among the two CHD studies performed to date, one reported that low IQ was associated with reduced CT in the left precuneus and the right caudal middle frontal cortex.^32^ The other identified reduced SA in the lateral sulcus is connected with worse executive function and increased CT in a wide range of regions linked with worse outcomes.^30^ However, few studies have examined CT in combination with SA or cortical volume in populations with CHD.

This paper aims to scrutinise regional cortical alterations in adolescents with CHD, and to assess how CT and SA contribute to cortical volume alterations. Furthermore, we will investigate how individual cortical structures are linked to cognitive functions. We hypothesise a widespread significant reduction in both CT and SA in individuals with CHD, but we can infer that alterations in cortical volume are mostly driven by SA rather than CT, since in healthy populations, individual differences in cortical volume are primarily attributed to variability in SA, not CT.^33^ We also expect to see strong associations between CT/SA and cognitive functions, as previous studies on structure-function relationships in healthy populations have consistently shown a positive association between SA and cognition, as well as age-dependent associations between CT and cognition.^34^

## Methods

### Participants

This analysis is part of a broader cohort study (TeenHeart Study^35^). Data were gathered at the University Children’s Hospital of Zurich from April 2019 to September 2021. The recruitment procedure is outlined in the aforementioned study protocol.

Adolescents with complex CHD were eligible for this study if they had undergone cardiopulmonary bypass (CPB) surgery between 2004 and 2012 at the University Children’s Hospital Zurich. Additional inclusion criteria were: (1) undergoing CPB surgery in the first year, (2) no diagnosis of a genetic or dysmorphic syndrome, and (3) being aged between 10 and 15 years at the time of evaluation. A total of 100 adolescents with CHD participated in the study (56% participation rate) from 178 eligible candidates. Additionally, 104 healthy adolescents aged 10 to 15 years were recruited as a control group. Exclusion criteria for the control group included: (1) birth before 36 weeks of gestation and (2) diagnosis of a neurological or significant developmental disorder (e.g., learning disability or attention deficit hyperactivity disorder).

Parental education was measured using a 6-point Likert scale (1 = no high school degree, 2 = high school degree, 3 = apprenticeship, 4 = higher diploma for craftsmen/craftswomen, 5 = advanced diploma of higher education, 6 = university degree). Socioeconomic status (SES) was derived by summing the highest education level of both parents, with possible scores ranging from 2 to 12.^36^ Clinical risk factors of patients were acquired from their clinical records, such as the length of intensive care unit (ICU) stay, surgery-related parameters, and history of stroke/seizure.

Ethical approval was obtained from the ethics committee of the Canton of Zurich. Written informed consent was secured from the legal guardians of all participants and the participants themselves if they were 14 years of age or older.

### Neuropsychological Assessment

#### Intelligence Quotient (IQ)

IQ was assessed with a corrected short version of the Wechsler Intelligence Scale for Children 4th edition (WISC-IV). This short version included the subtests *Matrices, Similarities, Letter Number Sequencing* and *Symbol Search*.^37,38^

#### Executive Functions (EF)

A comprehensive neuropsychological test battery was used to assess various domains of executive functions (EF), including working memory, inhibition, cognitive flexibility, planning, and fluency.^35^ The Delis-Kaplan Executive Function System,^39^ the Test of Attentional Performance (TAP),^40^ and the Regensburger Verbal Fluency Test^41^ were employed. Detailed descriptions of neuropsychological test measures of EF can be found in Supplementary Table 1. A summary score for overall EF performance was derived using the control group as the normative reference as previously described.^35,42^ To minimise the effects of fatigue and reduced motivation, tests were administered in a randomised order. The assessments were conducted and interpreted by trained psychologists and paediatricians.

### MRI acquisition

Cerebral MRI data were collected using a 3T GE MR750 scanner. Suitability for MRI acquisition was assessed prior to study participation using a safety screening questionnaire, which was filled out by the parents. All surgery reports were screened for ferromagnetic implants to confirm MR safety. For the MRI scan, hearing protection (earplugs and headsets) was provided.

High-resolution three-dimensional T1-weighted images were acquired using a three-dimensional spoiled gradient echo pulse sequence (SPGR) to assess brain volumes, cortical thickness and the presence of macroscopic lesions or other abnormalities. SPGR images were acquired using the following parameters: repetition time/echo time (TR/TE)=11/5 ms; inversion time=600 ms; flip angle=8°; reconstructed matrix=256 × 256; field of view (FOV)=26 cm; 176 contiguous axial slices, 1 mm slice thickness.

### MRI data preprocessing

To perform cortical reconstruction and volumetric segmentation of the whole brain^43,44^, the Freesurfer image analysis suite (version 7.1, http://surfer.nmr.mgh.harvard.edu) was used. The automated recon-all pipeline consists of more than 30 stages, including motion correction, intensity normalisation, a Talairach transform of each subject’s native brain, removal of the nonbrain tissue, segmentation of the subcortical grey matter/white matter (GM/WM) tissue and GM/WM boundary tessellation. The quality of the FreeSurfer segmentation was visually inspected by AT and RT, and scans with poor image quality or suboptimal segmentation were excluded, but images with subtle motion artefacts were retained. Images from patients were particularly inspected to ensure that they had no abnormalities that could affect subsequent analysis.

Typically, each participant’s brain is registered to a standard template like fsaverage based on the gyral and sulcal pattern. This standardisation allows for a comparison of cortical metrics across participants in a common space.^45^ Instead of using the fsaverage template, which is created based on adult brains, we created a custom group-average template to better represent the anatomical features of children and adolescents, including patients with CHD. The instructions for initialising the custom template with the FreeSurfer template can be found on this website (https://surfer.nmr.mgh.harvard.edu/fswiki/SurfaceRegAndTemplates). Each participant’s cortical surfaces were then registered to this custom group-average template, ensuring a more accurate alignment of cortical features across participants.

After registration to the template, CT and SA values were extracted for each participant. CT was measured as the shortest distance between the pial surface and the GM/WM boundary, and the mean CT for each participant was computed by averaging the CT at each vertex for both hemispheres. SA was calculated as the total area of the cortical mantle on the pial surface. The entire cerebral cortex was then parcellated into 34 distinct regions per hemisphere using the Desikan-Killiany atlas,^46^ to create maps of volume, CT and SA. These cortical maps were smoothed using a 10 mm full-width at half-maximum (FWHM) surface-based Gaussian kernel to lower local anatomical variation and facilitate group-level statistical analysis. It is reported that within-scanner variabilities of MRI-derived metrics using Freesurfer were negligible.^47,48^

### Statistical analysis

#### General statistical analysis

Statistical analyses of the whole-brain mean CT, total SA, and cortical volume were carried out using R software,^49^ version 4.3.2. Descriptive statistics comprised mean (standard deviation: SD) and median (interquartile range: IQR) for continuous variables and counts and proportions for categorical ones. Groups (such as the CHD and control groups) were compared using a two-sample t-test for continuous variables (e.g., age), a Mann-Whitney test for ordinal variables (e.g., SES) or a Chi-squared test for categorical ones (e.g., sex). Missing SES data (12 out of 100 in the CHD group and 12 out of 104 in the control group) were handled using group-wise median imputation. To assess potential bias, the subset of individuals without imaging data or those excluded due to artefacts was compared with the final sample demographics.

#### Statistical analysis for total/mean cortical structures

Statistical analyses of the whole-brain mean CT, total SA, and cortical volume were controlled for age, sex, and SES. The interrelationships between cortical metrics were investigated using a Pearson’s correlation matrix. Multiple linear regression was used to test if total cortical volumes are associated with total SA and mean CT (cortical volume ∼ total SA + mean CT + group + age + sex + SES). To compare the predictive strength of the total SA and the mean CT in explaining cortical volume, we employed a bootstrapping approach. This analysis estimated the sampling distribution of standardised slopes (beta coefficients) 1,000 times and their 95% confidence intervals (CIs) using the bias-corrected and accelerated method. By examining the overlap and width of these CIs, we determined whether the strength of association between cortical volume and SA differs significantly from that between cortical volume and CT.

CHD-control group differences and group differences among further CHD subgroups (univentricular or biventricular; cyanotic or acyanotic) in cortical metrics were evaluated using multiple linear regression. Interaction effects (group:age, group:sex, and group:SES) on cortical metrics were analysed through a multiple linear regression. Additionally, in the CHD group, an association between cortical metrics and the length of ICU stay was investigated, as this clinical risk factor has previously been identified as a predictive marker for neurodevelopmental outcomes.^50^

Furthermore, associations between cognitive outcomes (IQ and EF summary score; dependent variables) and cortical metrics (independent variables) were calculated using multiple linear regression. Interaction effects (cortical metrics:group) were tested to investigate if the strength of association between cortical metrics and cognitive outcomes differs between patients and controls. All the P-values for linear regressions were adjusted for multiple comparisons using the false discovery rate (FDR). Statistical significance was demarcated at a two-tailed p < 0.05.

#### MRI-based statistical analysis (GLM)

Group differences (CHD and controls) in regional-level cortical volumes, SA, and CT were investigated using vertex-based multivariate General Linear Model (GLM) analysis, controlling for age and sex. A FreeSurfer-programmed ‘Different Onset, Different Slope’ (DODS) GLM was applied to the spatially normalised cortical data to create statistical maps depicting the group differences in cortical metrics, regressing out the effects of age and sex. Additionally, we covaried for the total SA and mean CT separately to see if there are any disproportionately smaller or thinner regions irrespective of the cortex size/thickness for the CHD group.

To examine the cortical regions associated with IQ and EF across the entire sample, the same DODS model was used, incorporating additional IQ/EF covariates while adjusting for age, sex, and group. Thereafter, interaction effects were investigated to see if there were any group differences in the strength and direction of association between cortical metrics and IQ/EF.

Monte Carlo simulation data, necessary for correcting multiple comparisons, was precomputed using 10,000 iterations to ensure an accurate estimation of the null distribution of cluster sizes. Then, cortical surface results were corrected for multiple comparisons using Freesurfer’s cluster-wise correction, with a vertex-wise cluster-forming threshold of p < 0.05 (two-tailed), and adjusted for two hemispheres using the Bonferroni correction. The results with a 10 mm FWHM smoothing value were reported based on a previous report regarding clusterwise false positive rates when applying different FWHM values after the permutation simulation correction.^51^ As an additional validation approach, we re-ran all GLM analyses in a subsample with best image quality only (without any minor movement artefacts, N=111).

## Results

### Demographic data

Of the 204 participants who participated in the study (100 patients, 104 controls), 155 underwent cerebral MRI (60 patients, 95 controls). The main reasons for not undergoing cerebral MRI were implants in patients with CHD (e.g., pacemaker, stents, clips), or anxiety. Among those with MRI data, 26 (11 patients, 15 controls) were excluded due to poor image quality or suboptimal segmentation. Macroscopic brain abnormalities were reviewed, but did not affect cortical segmentation and analysis. Thus, the final sample size is N=129 (49 patients, 80 controls) with a mean age of 13.2 years (SD=1.3). The CHD group was older than the control group (t(df)=-3.56(117), p=0.001). There was no difference in sex distribution between groups (Χ2(df)=2.03(1), p=0.154). The CHD group had lower SES (W=1006.5, p<0.001), lower IQ (t(df)= −5.63(86), p<0.001) and lower EF (t(df)= −4.83(95), p<0.001) than controls. The sample characteristics of patients and controls are presented in Table 1, and the CHD diagnoses are listed in Supplementary Table 2.

**Table 1:** Demographic data of participants.

| Sample characteristics | CHD (n=49) | Controls (n=80) | p-value |
| --- | --- | --- | --- |
| Age, mean (SD) | 13.65 (1.1) | 12.86 (1.4) | 0.001 |
| Male sex, N (%) | 31 (63) | 39 (49) | 0.154<sup>a</sup> |
| SES, median (IQR) | 7 (6-9) | 10 (8-12) | < 0.001<sup>b</sup> |
| Gestational age, mean (SD), wk | 39.40 (1.6) | 39.06 (1.5) | 0.310 |
| Birth weight, mean (SD), g | 3295 (559.9) | 3174 (539.3) | 0.580 |
| IQ, mean (SD) | 99.69 (11.6) | 110.96 (9.7) | < 0.001 |
| EF summary score, mean (SD), z | -0.99 (1.1) | -0.06 (1.0) | < 0.001 |
| **CHD characteristics** |  |  |  |
| Prenatal diagnosis, N (%) | 9 (18) | - |  |
| Univentricular CHD, N (%) | 9 (18) | - |  |
| Cyanotic CHD, N (%) | 34 (69) | - |  |
| **Perioperative time of first CPB surgery** |  |  |  |
| Preoperative O<sub>2</sub> saturation, mean (SD), % | 87.17 (10) | - |  |
| Age at surgery, mean (SD), month | 2.58 (2.9) | - |  |
| Lowest intraoperative temperature, mean (SD), °C | 28.94 (3.9) | - |  |
| ECC time, mean (SD), min | 170.27 (72.1) | - |  |
| Length of hospital stay, median (IQR), days | 21.00 (17-36) | - |  |
| Length of ICU stay, median (IQR), days | 7.00 (5-10) | - |  |
| No. CPB surgeries, median (IQR) | 1 (1-3) | - |  |
| **Critical neurological or cardiac events ever** |  |  |  |
| ECMO, N (%) | 1 (2) | - |  |
| Clinically relevant seizure, N (%) | 2 (4) | - |  |
| Stroke identified on cerebral MRI, N (%) | 4 (8) | - |  |
| Number of cardiac medications* at assessment, median (IQR) | 0 (0-4) | - |  |

Note: CHD = congenital heart disease, ECC = extracorporeal circulation, ECMO = extracorporeal membrane oxygenation, ICU = intensive care unit, CPB = cardiopulmonary bypass. SD = standard deviation, IQR = interquartile range.
<sup>a</sup> Two-sampled $\chi^2$ test.
<sup>b</sup> Two-sampled Mann-Whitney U test.
Others: Two-sampled independent t-test.
\* cardiac medications are mainly ACE inhibitor, Beta Blocker, Diuretic, Aspirin.

The final sample did not differ from excluded participants (N=75: 51 patients, 24 controls) regarding age (t(df)=-1.56(154), p=0.121), sex (Χ2(df)=0.00(1), p=1.00), SES (W=5594, p=0.060), IQ (t(df)= 1.87(110), p= 0.064), the proportion of cyanotic CHD (Χ2(df)=0.33(1), p= 0.567) or univentricle CHD (Χ2(df)=1.12(1), p= 0.290) and the length of ICU stay for patients (t(df)=-1.96(51.7), p=0.055).

### Total surface area and mean cortical thickness

Correlations between total SA and mean CT were weak for both hemispheres (left: r=0.26, right: r=0.29). Across the whole sample, total cortical volume correlated strongly with total SA, but only weakly with mean CT (i.e., volume ∼ total SA: left β =0.868, right β =0.857; volume ∼ mean CT: left β =0.281, right β =0.298). Figure 1 shows the difference in correlation strength of the total SA and cortical volume, mean CT and cortical volume with different R^2^ model fits (volume ∼ total SA: left R^2^ =0.910, right R^2^ =0.910; volume ∼ mean CT: left R^2^ =0.248, right R^2^ =0.291). Furthermore, bootstrapping revealed that SA has a stronger and more consistent relationship with cortical volume (95_CI left: [0.936, 0.966], right: [0.933, 0.966]) compared to CT (95_CI left: [0.359, 0.624], right: [0.407, 0.656]), as evidenced by the higher and narrower confidence interval for SA. Moreover, the lack of overlap between the two intervals suggests a potential difference in the predictive strength of these two variables.

**Figure 1.**
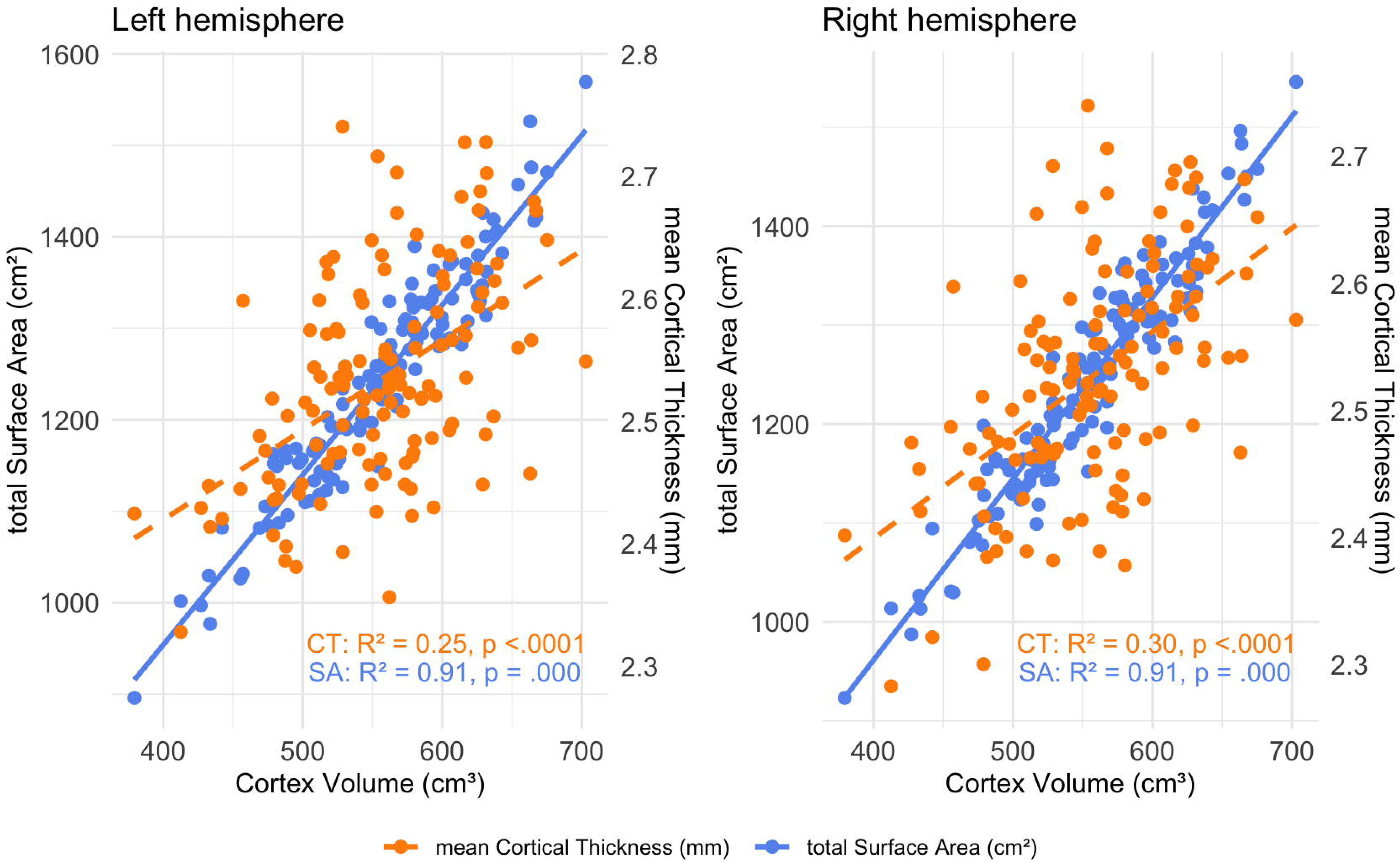
Comparison of the correlation strength of the surface area and cortical volume, cortical thickness and cortical volume. Orange dots with dash line: mean cortical thickness (CT), blue dots with solid line: total surface area (SA). This dual y-axis plot shows that the correlation strength of SA (primary y-axis) and cortical volume (x-axis) is stronger than CT (second y-axis) and cortical volume. Left hemisphere on the left and right hemisphere on the right.

Patients with CHD showed statistically significantly lower values in total/mean cortical metrics (i.e., cortical volume, total SA, mean CT in both hemispheres) compared to controls (see Figure 2) after controlling for covariates and FDR correction. Statistical details are provided in Supplemental Table 3. Furthermore, there were no statistically significant interaction effects of group:age, group:sex and group:SES on any cortical metrics, except for group:SES effects on total SA (left: β = −0.754, p=0.036, right: β = −0.789, p=0.034), indicating the effect of SES on total SA (but not volume or mean CT) is weaker in controls than the CHD group.

**Figure 2.**
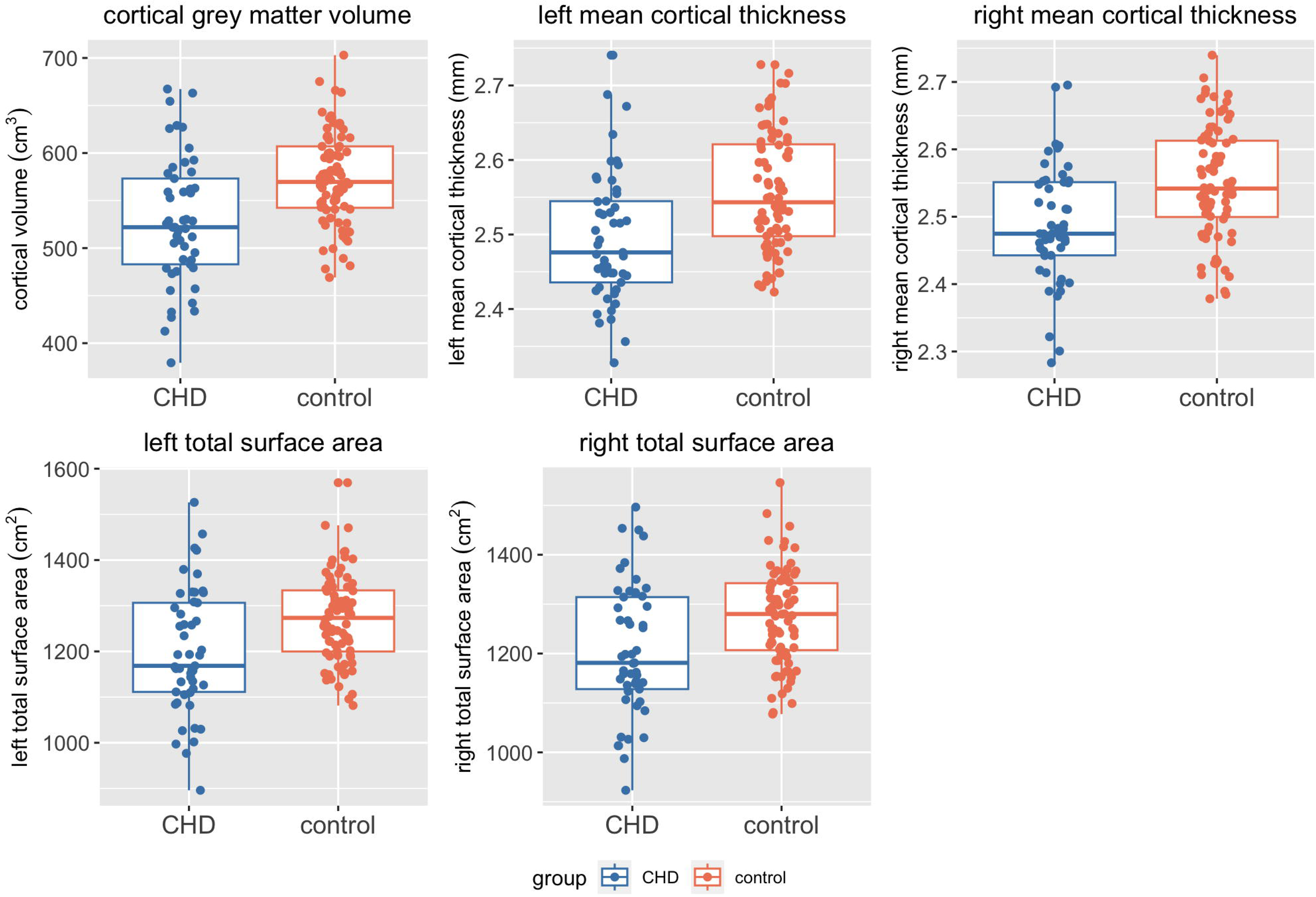
Comparison of the CHD and control group in cortical volume, mean cortical thickness and total surface area. Blue = CHD, Red = Control. The CHD group showed lower values in cortical volume, mean cortical thickness and total surface area in both hemispheres.

Among the CHD group, patients with cyanotic CHD showed lower mean CT than patients with acyanotic CHD (left | β = 0.327, 95_CI[0.052, 0.601], p = 0.049; right | β = 0.399, 95_CI[0.133, 0.664], p = 0.017). Similarly, patients with univentricular defects showed lower mean CT than those with biventricular defects (left | β = 0.438, 95_CI [0.200, 0.675], p = 0.003; right | β = 0.492, 95_CI [0.264, 0.720], p = 0.001). However, no statistically significant difference was observed in cortical volume and total SA between any of these subgroups. Furthermore, the length of ICU stay after the first CPB surgery was not associated with any cortical metrics (cortical volume: β = −0.087, left mean CT: β = −0.096, right mean CT: β = −0.063, left total SA: β = −0.029, right total SA: β = −0.047).

### Regional cortical reductions in patients with CHD

After confirming the global reduction in cortical metrics (total SA, mean CT, and cortical volume) in patients with CHD, we conducted a Freesurfer regional analysis to identify brain areas that are particularly affected in these patients. Comparisons of cortical SA, CT and volume between patients with CHD and healthy controls (adjusted for age and sex) are shown in Figure 3A. After cluster-wise multiple testing corrections, the CHD group showed significantly lower SA in 5 clusters (average cluster area size = 163.88 cm^2^), lower CT in 7 small clusters (average cluster area size = 25.26 cm^2^) and lower cortical volume in 11 clusters (average cluster area size = 47.88 cm^2^) bilaterally. The cluster statistics are presented in Table 2. For supplementary purposes to confirm imaging findings, the main results were compared with the selected subsample (N=111) that had no minor movement artefacts; however, the results remained largely unchanged (See Supplemental Figure 1).

**Figure 3.**
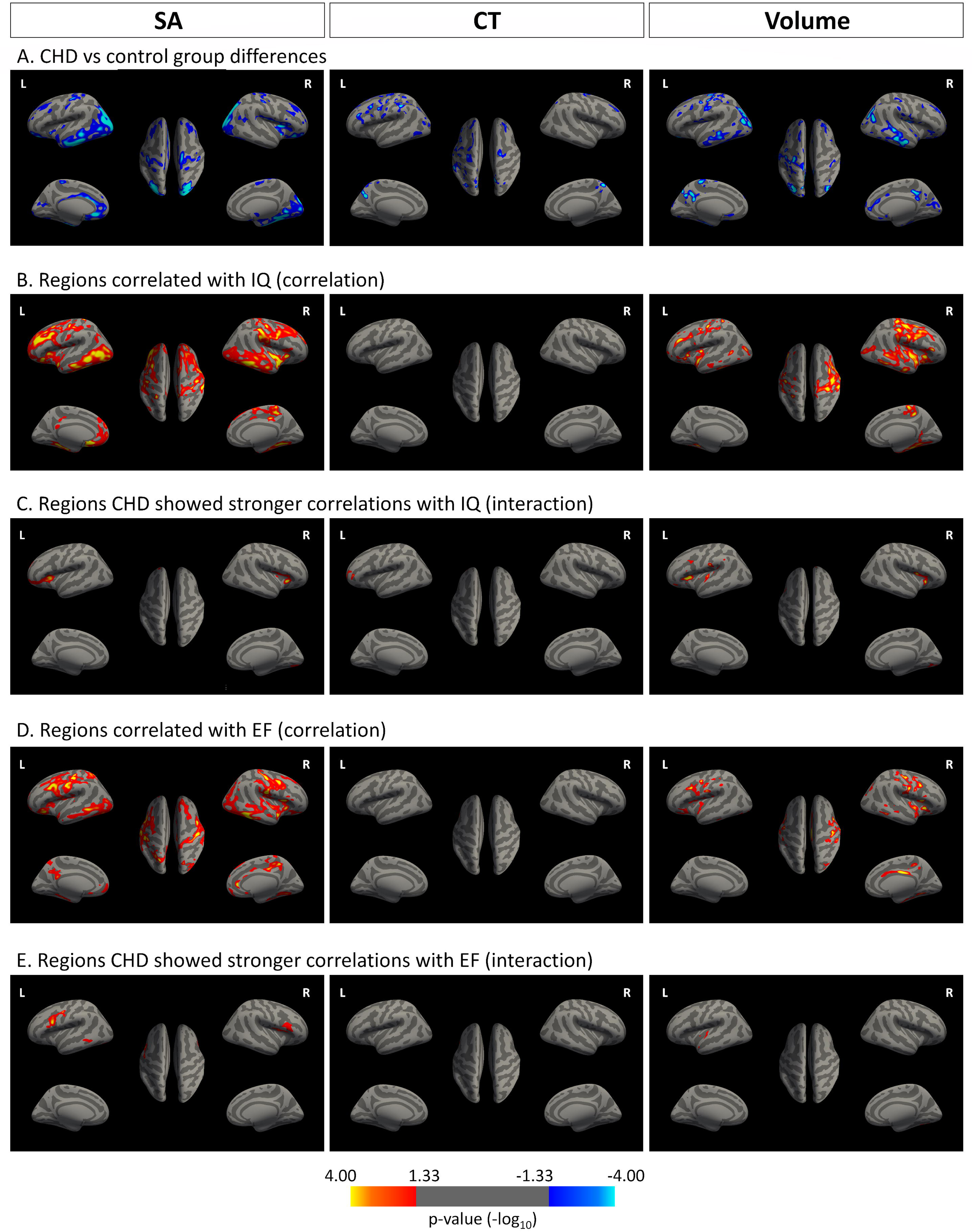
(A∼E) cortical surface structures, comparison between groups, association with IQ/EF summary score. All analyses were adjusted for age and sex, p < 0.05, cluster-wise corrected, represented by blue clusters (negative correlation) or yellow-red clusters (positive correlation). The colour bar represents uncorrected significance values masked by the clusters that survived correction for multiple comparisons. SA = surface area, CT = cortical thickness, Volume = cortical volume. EF = executive function **A.** Patients with CHD showed significantly lower SA than controls in the left inferior parietal and postcentral, the right middle temporal and rostral medial frontal and supramarginal regions (the leftmost), significantly lower CT in the bilateral precuneus, precentral and rostral middle frontal, the left inferior temporal region (the middle figure), and significantly lower volume in the left superior parietal, inferior temporal, precuneus, caudal middle frontal, lateral orbitofrontal, superior frontal, and the right inferior parietal, lateral orbitofrontal, posterior cingulate and postcentral regions (the rightmost). No significant positive clusters (yellow-red) were found. **B.** SA in the left medial orbitofrontal, isthmus cingulate and the right superior temporal regions were associated with IQ (the leftmost). No significant clusters associated with IQ were found for CT (the middle). For cortical volume, the bilateral fusiform, superior temporal, and lateral occipital, the left rostral middle frontal, and the right postcentral, lateral orbitofraontal and paracentral regions were associated with IQ (the rightmost). No significant negative clusters (blue) were found. **C.** The CHD group showed stronger associations between IQ and SA in the left pars triangularis, lingual and the right lateral orbitofrontal regions (the leftmost). For CT, the left rostral middle frontal region showed a stronger correlation with IQ in the CHD than the control group (middle). For volume, the left supramarginal, pars triangularis, caudal middle frontal and the right lateral orbitofrontal and lingual regions showed stronger associations with IQ (the rightmost) **D.** SA in the left postcentral and the right pars opercularis regions were associated with the EF summary score (the leftmost). No significant clusters associated with EF summary score were found for CT (the middle). For cortical volume, the bilateral postcentral, the left caudal middle frontal and fusiform, the right pars opercularis, posterior cingulate, lingual, inferior temporal, inferior parietal regions were associated with the EF summary score (the rightmost). No significant negative clusters (blue) were found. **E.** The CHD group showed stronger associations between EF summary score and SA in the bilateral pars opercularis, the left lateral occipital regions (the leftmost). For CT, there were no clusters where the group difference in EF summary score was significant (middle). For volume, the left superior temporal region showed stronger associations with EF summary score than controls (the rightmost)

**Table 2:** Cortical clusters comparison between the CHD and control group.

| Annotation | side | Max | NVtx | cluster size<br>(cm<sup>2</sup>) | clusterwise<br>p-value | Talairach coordinates |  |  |
| --- | --- | --- | --- | --- | --- | --- | --- | --- |
|  |  |  |  |  |  | X | Y | Z |
| Surface area |  |  |  |  |  |  |  |  |
| inferior parietal | L | -8.27 | 126268 | 373.81 | 0.0002 | -30 | -74.6 | 18.8 |
| postcentral | L | -4.09 | 92538 | 50.78 | 0.0002 | -40 | -28 | 49.3 |
| middle temporal | R | -6.00 | 105454 | 238.57 | 0.0002 | 59.2 | -21.6 | -18.2 |
| rostral middle frontal | R | -4.50 | 121751 | 134.62 | 0.0002 | 19.6 | 56 | 22.6 |
| supramarginal | R | -3.09 | 98724 | 21.60 | 0.0056 | 49.5 | -27.1 | 38.2 |
| Cortical thickness |  |  |  |  |  |  |  |  |
| precuneus | L | -6.59 | 13896 | 65.78 | 0.0002 | -6 | -71.6 | 30 |
| precentral | L | -4.01 | 129230 | 46.26 | 0.0002 | -56 | -1.5 | 37.9 |
| rostral middle frontal | L | -4.09 | 48842 | 20.34 | 0.0002 | -43.6 | 33.4 | 24.8 |
| inferior temporal | L | -3.16 | 17450 | 8.60 | 0.0365 | -49.9 | -64.5 | -4 |
| precuneus | R | -6.31 | 71304 | 17.55 | 0.0002 | 10.3 | -60.5 | 53 |
| precentral | R | -3.82 | 137882 | 9.43 | 0.0197 | 19.6 | -17.2 | 62.5 |
| rostral middle frontal | R | -3.69 | 123206 | 8.83 | 0.0316 | 30.2 | 35.3 | 31.9 |
| Cortical volume |  |  |  |  |  |  |  |  |
| superior parietal | L | -6.67 | 157742 | 125.72 | 0.0002 | -28.4 | -72 | 20.5 |
| inferior temporal | L | -5.89 | 7115 | 81.66 | 0.0002 | -45.3 | -30.5 | -20.6 |
| precuneus | L | -5.39 | 136087 | 22.80 | 0.0002 | -13.5 | -55.2 | 33.9 |
| caudal middle frontal | L | -3.41 | 123129 | 16.51 | 0.0002 | -34.5 | 4.9 | 35.9 |
| lateral orbitofrontal | L | -5.64 | 67784 | 14.10 | 0.0004 | -22.3 | 36.2 | -11.8 |
| superior frontal | L | -3.80 | 65524 | 12.60 | 0.0020 | -13.9 | 52.2 | 31 |
| inferior parietal | R | -5.77 | 147528 | 103.23 | 0.0002 | 35.6 | -74.3 | 34 |
| lateral orbitofrontal | R | -3.72 | 50935 | 71.62 | 0.0002 | 15.2 | 31.7 | -23.3 |
| posterior cingulate | R | -4.10 | 141645 | 38.44 | 0.0002 | 13.4 | -38.3 | 38.3 |
| postcentral | R | -4.98 | 98756 | 25.59 | 0.0002 | 38.1 | -13.6 | 22.4 |
| postcentral | R | -2.90 | 140326 | 14.35 | 0.0004 | 41 | -33.8 | 55.7 |

p < 0.05, corrected for multiple comparisons; MAX = the maximum -log10(p-value) in the cluster, NVtx = the vertex number at the maximum.

As an additional analysis to examine the regional specificity of these findings, we covaried for the total SA to see if there are any disproportionately smaller regions irrespective of the cortex size for the CHD group. We found that the bilateral inferior temporal SA (left area size = 27.02cm^2^, right = 20.19cm^2^) and the right inferior parietal SA (area size = 25.75cm^2^) were particularly smaller in patients with CHD compared to controls when total SA size was accounted for. For CT, there were no regions where group differences were significant when accounting for the mean CT.

### Cognitive outcomes and cortical structures

On the global level, IQ was significantly associated with all the cortical metrics (mean CT showing the weakest association) across the whole sample. In contrast to IQ, the EF was associated only with cortical volume and total SA, not with mean CT. Statistical details are presented in Table 3. The interaction effect was non-significant, indicating that the strength of associations between IQ/EF and cortical metrics did not differ between groups.

**Table 3:** Multiple regression for IQ/EF summary score and cortical metrics across the whole sample (N=129)

| Effect | B | Standard error | 95% CI | $\beta$ | p-value | R<sup>2</sup> | p-model fit |
| --- | --- | --- | --- | --- | --- | --- | --- |
| IQ ~ Cortical volume |  |  |  |  |  |  |  |
| cortical volume | 0.084 | 0.018 | 0.257 ~ 0.590 | 0.423 | 1.06E-05 | 0.351 | 1.39E-11 |
| age | 0.463 | 0.687 | -0.096 ~ 0.199 | 0.051 | 0.686 |  |  |
| sex | 4.027 | 1.939 | 0.012 ~ 0.330 | 0.171 | 0.066 |  |  |
| SES | 1.881 | 0.424 | 0.207 ~ 0.504 | 0.356 | 2.24E-05 |  |  |
| IQ ~ lh mean CT |  |  |  |  |  |  |  |
| lh mean CT | 29.454 | 11.236 | 0.059 ~ 0.377 | 0.218 | 0.011 | 0.273 | 1.10E-08 |
| age | 0.460 | 0.765 | -0.113 ~ 0.215 | 0.051 | 0.686 |  |  |
| sex | 0.173 | 1.822 | -0.143 ~ 0.157 | 0.007 | 0.924 |  |  |
| SES | 2.675 | 0.415 | 0.372 ~ 0.639 | 0.505 | 4.23E-09 |  |  |
| IQ ~ rh mean CT |  |  |  |  |  |  |  |
| rh mean CT | 28.504 | 11.301 | 0.052 ~ 0.383 | 0.218 | 0.013 | 0.270 | 1.39E-08 |
| age | 0.538 | 0.783 | -0.109 ~ 0.228 | 0.060 | 0.686 |  |  |
| sex | 0.372 | 1.835 | -0.135 ~ 0.167 | 0.016 | 0.924 |  |  |
| SES | 2.594 | 0.416 | 0.355 ~ 0.626 | 0.490 | 1.00E-08 |  |  |
| IQ ~ lh total SA |  |  |  |  |  |  |  |
| lh total SA | 0.040 | 0.009 | 0.219 ~ 0.558 | 0.388 | 5.34E-05 | 0.333 | 7.27E-11 |
| age | 0.170 | 0.686 | -0.129 ~ 0.166 | 0.019 | 0.895 |  |  |
| sex | 3.827 | 1.982 | 0.000 ~ 0.325 | 0.162 | 0.080 |  |  |
| SES | 1.887 | 0.435 | 0.204 ~ 0.509 | 0.357 | 3.00E-05 |  |  |
| IQ ~ rh total SA |  |  |  |  |  |  |  |
| rh total SA | 0.037 | 0.009 | 0.189 ~ 0.529 | 0.359 | 1.57E-04 | 0.320 | 2.22E-10 |
| age | 0.072 | 0.690 | -0.141 ~ 0.157 | 0.008 | 0.917 |  |  |
| sex | 3.587 | 2.004 | -0.012 ~ 0.317 | 0.152 | 0.095 |  |  |
| SES | 1.974 | 0.435 | 0.221 ~ 0.525 | 0.373 | 1.72E-05 |  |  |
| EF ~ Cortical volume |  |  |  |  |  |  |  |
| cortical volume | 0.006 | 0.002 | 0.154 ~ 0.527 | 0.340 | 0.001 | 0.208 | 1.14E-06 |
| age | 0.035 | 0.070 | -0.120 ~ 0.202 | 0.041 | 0.894 |  |  |
| sex | 0.319 | 0.203 | -0.032 ~ 0.316 | 0.142 | 0.168 |  |  |
| SES | 0.135 | 0.044 | 0.103 ~ 0.438 | 0.271 | 0.003 |  |  |
| EF ~ lh mean CT |  |  |  |  |  |  |  |
| lh mean CT | 1.422 | 1.146 | -0.062 ~ 0.283 | 0.111 | 0.217 | 0.144 | 1.08E-04 |
| age | 0.014 | 0.076 | -0.158 ~ 0.193 | 0.017 | 0.894 |  |  |
| sex | 0.010 | 0.186 | -0.156 ~ 0.165 | 0.004 | 0.957 |  |  |
| SES | 0.197 | 0.042 | 0.243 ~ 0.545 | 0.394 | 1.19E-05 |  |  |
| EF ~ rh mean CT |  |  |  |  |  |  |  |
| rh mean CT | 1.467 | 1.158 | -0.062 ~ 0.296 | 0.117 | 0.217 | 0.144 | 1.04E-04 |
| age | 0.022 | 0.078 | -0.155 ~ 0.206 | 0.026 | 0.894 |  |  |
| sex | 0.019 | 0.187 | -0.153 ~ 0.170 | 0.008 | 0.957 |  |  |
| SES | 0.193 | 0.042 | 0.236 ~ 0.539 | 0.388 | 1.47E-05 |  |  |
| EF ~ lh total SA |  |  |  |  |  |  |  |
| lh total SA | 0.003 | 0.001 | 0.139 ~ 0.514 | 0.327 | 0.002 | 0.202 | 1.73E-06 |
| age | 0.015 | 0.069 | -0.141 ~ 0.177 | 0.018 | 0.894 |  |  |
| sex | 0.317 | 0.205 | -0.035 ~ 0.318 | 0.141 | 0.168 |  |  |
| SES | 0.134 | 0.045 | 0.099 ~ 0.438 | 0.269 | 0.003 |  |  |
| EF ~ rh total SA |  |  |  |  |  |  |  |
| rh total SA | 0.003 | 0.001 | 0.129 ~ 0.502 | 0.315 | 0.002 | 0.199 | 2.24E-06 |
| age | 0.009 | 0.069 | -0.148 ~ 0.170 | 0.011 | 0.894 |  |  |
| sex | 0.310 | 0.206 | -0.039 ~ 0.315 | 0.138 | 0.168 |  |  |
| SES | 0.138 | 0.045 | 0.109 ~ 0.445 | 0.277 | 0.003 |  |  |

CT = cortical thickness (mm), SA = surface area (cm<sup>2</sup>), lh = left hemisphere, rh = right hemisphere, CI = confidence interval, p-values are FDR corrected. EF = executive function

On the regional level, statistically significant positive associations were found between IQ and SA, as well as cortical volume, across the entire sample, but not with CT (See Figure 3B). The association between IQ and SA was especially strong in the left medial orbitofrontal and right superior temporal regions. Furthermore, there were several regions where the CHD group showed a stronger association with IQ in every cortical metric (See Figure 3C). Interestingly, one CT cluster in the rostral middle frontal region remained significant in the group-IQ interaction analysis, indicating that the CHD group had stronger relationships between CT and IQ in this region, despite the absence of a CT-IQ association in the full sample. There were also several regions of SA that showed stronger associations with IQ in the CHD group than the control group. Cluster statistics are presented in Table 4.

**Table 4:** Cortical clusters statistics, association and interaction with IQ.

| Cortical clusters that are associated with IQ |  |  |  |  |  | Talairach coordinates |  |  |
| --- | --- | --- | --- | --- | --- | --- | --- | --- |
| Annotation | side | Max | NVtx | cluster size (cm2) | clusterwise p-value | X | Y | Z |
| **Surface area** |  |  |  |  |  |  |  |  |
| medial orbitofrontal | L | 5.77 | 154316 | 472.12 | 0.0002 | -8 | 19.9 | -11.5 |
| isthmus cingulate | L | 2.95 | 94473 | 16.35 | 0.04723 | -5.7 | -45.8 | 22 |
| superior temporal | R | 5.84 | 44098 | 426.95 | 0.0002 | 47.1 | -16.5 | -2.8 |
| **Cortical thickness** |  |  |  |  |  |  |  |  |
| No significant clusters were found. |  |  |  |  |  |  |  |  |
| **Cortical volume** |  |  |  |  |  |  |  |  |
| rostral middle frontal | L | 5.90 | 18121 | 62.55 | 0.0002 | -44.2 | 31.7 | 25.7 |
| lateral occipital | L | 3.82 | 45886 | 50.42 | 0.0002 | -40.7 | -80.9 | 9.4 |
| superior temporal | L | 4.88 | 79243 | 48.83 | 0.0002 | -45.9 | -6.2 | -12.2 |
| fusiform | L | 5.02 | 163034 | 13.87 | 0.0006 | -35.7 | -42.2 | -11.1 |
| rostral middle frontal | L | 2.48 | 55082 | 10.38 | 0.01017 | -27.1 | 30.7 | 35.7 |
| postcentral | R | 5.66 | 64037 | 74.56 | 0.0002 | 32.2 | -32.4 | 57.5 |
| superior temporal | R | 4.99 | 63175 | 50.10 | 0.0002 | 45.5 | -16.9 | -4.7 |
| fusiform | R | 4.93 | 48225 | 46.89 | 0.0002 | 36.4 | -36.9 | -17.4 |
| lateral orbitofrontal | R | 5.79 | 1141 | 13.56 | 0.0012 | 29.2 | 24.2 | 0.9 |
| lateral occipital | R | 2.61 | 129166 | 9.65 | 0.01653 | 23.8 | -100.8 | 1.4 |
| paracentral | R | 4.92 | 150120 | 8.65 | 0.03666 | 18.4 | -42.8 | 45.1 |
| **Cortical clusters that had a stronger association with IQ in CHD than controls** |  |  |  |  |  |  |  |  |
|  |  |  |  |  |  | Talairach coordinates |  |  |
| Annotation | side | Max | NVtx | cluster size (cm2) | clusterwise p-value | X | Y | Z |
| **Surface area** |  |  |  |  |  |  |  |  |
| pars triangularis | L | 3.88 | 16896 | 54.10 | 0.0002 | -29.2 | 22.4 | 13.7 |
| lingual | R | 3.08 | 31904 | 19.94 | 0.00679 | 16.6 | -89.2 | -9.1 |
| lateral orbitofrontal | R | 4.89 | 27185 | 19.36 | 0.00838 | 29.2 | 23.3 | 1.9 |
| **Cortical thickness** |  |  |  |  |  |  |  |  |
| rostral middle frontal | L | 3.57 | 141255 | 13.19 | 0.0012 | -38.8 | 42.3 | 4.2 |
| **Cortical volume** |  |  |  |  |  |  |  |  |
| supramarginal | L | 3.84 | 95048 | 18.15 | 0.0002 | -59 | -21.3 | 24.3 |
| pars triangularis | L | 5.50 | 38015 | 14.89 | 0.0002 | -28.2 | 25.8 | 10.6 |
| caudal middle frontal | L | 3.13 | 66571 | 8.84 | 0.03155 | -32.6 | 3.4 | 27.5 |
| lateral orbitofrontal | R | 4.54 | 51070 | 11.00 | 0.00579 | 29.1 | 23.8 | 0.5 |
| lingual | R | 2.66 | 70001 | 8.77 | 0.03292 | 15.8 | -90.7 | -8.1 |

p < 0.05, corrected for multiple comparisons; MAX = the maximum -log10(p-value) in the cluster, NVtx = the vertex number at the maximum.

The EF was significantly associated with regional SA and cortical volume, but not with regional CT across the whole sample (See Figure 3D). The association between the EF and SA was especially strong in the bilateral postcentral region. Significant interaction effects revealed that several regions in SA and volumes showed stronger associations with EF in patients with CHD than in controls. There were no significant CT clusters in this interaction analysis (See Figure 3E). Cluster statistics are described in Table 5.

**Table 5:** Cortical clusters statistics, association and interaction with EF summary score.

| Cortical clusters that are associated with EF summary score |  |  |  |  |  |  |  |  |
| --- | --- | --- | --- | --- | --- | --- | --- | --- |
| Annotation | side | Max | NVtx | cluster size (cm2) | clusterwise p-value | Talairach coordinates |  |  |
|  |  |  |  |  |  | X | Y | Z |
| Surface area |  |  |  |  |  |  |  |  |
| postcentral | L | 5.44 | 80831 | 376.00 | 0.0002 | -51.7 | -21 | 53.9 |
| pars opercularis | R | 5.49 | 103679 | 405.07 | 0.0002 | 41 | 14.4 | 11.2 |
| Cortical thickness |  |  |  |  |  |  |  |  |
| No significant clusters were found. |  |  |  |  |  |  |  |  |
| Cortical volume |  |  |  |  |  |  |  |  |
| caudal middle frontal | L | 4.63 | 66574 | 40.69 | 0.0002 | -31.7 | 4.8 | 28.2 |
| postcentral | L | 3.18 | 58528 | 29.80 | 0.0002 | -45.9 | -9.8 | 14.5 |
| fusiform | L | 2.72 | 90524 | 17.53 | 0.0002 | -39.8 | -77.2 | -15.5 |
| postcentral | R | 4.59 | 19998 | 54.10 | 0.0002 | 61 | -9.6 | 27.7 |
| pars opercularis | R | 5.19 | 72960 | 33.42 | 0.0002 | 39.9 | 14.5 | 12 |
| posterior cingulate | R | 5.77 | 146637 | 16.87 | 0.0002 | 4.5 | -13.9 | 31.5 |
| lingual | R | 3.71 | 3905 | 15.13 | 0.0004 | 28.1 | -57.6 | -4.3 |
| inferior parietal | R | 3.23 | 115812 | 13.53 | 0.0014 | 35.3 | -75.4 | 26.8 |
| inferior temporal | R | 3.72 | 114694 | 13.26 | 0.0014 | 59 | -49.1 | -15.2 |
| Cortical clusters that had a stronger association with EF summary score in CHD than controls |  |  |  |  |  |  |  |  |
| Annotation | side | Max | NVtx | cluster size (cm2) | clusterwise p-value | Talairach coordinates |  |  |
|  |  |  |  |  |  | X | Y | Z |
| Surface area |  |  |  |  |  |  |  |  |
| pars opercularis | L | 4.20 | 107997 | 25.17 | 0.0024 | -40.1 | 5.2 | 21.4 |
| lateral occipital | L | 2.56 | 59230 | 23.33 | 0.0042 | -33.2 | -86.4 | -15 |
| pars opercularis | R | 3.47 | 133950 | 21.10 | 0.00619 | 39.2 | 4.4 | 14.2 |
| Cortical thickness |  |  |  |  |  |  |  |  |
| No significant clusters were found. |  |  |  |  |  |  |  |  |
| Cortical volume |  |  |  |  |  |  |  |  |
| superior temporal | L | 2.57 | 125308 | 12.11 | 0.0032 | -53.4 | -12.2 | 3.4 |

p < 0.05, corrected for multiple comparisons; MAX = the maximum -log10(p-value) in the cluster, NVtx = the vertex number at the maximum.

## Discussion

In the present study, we examined the global and regional cortical structures of adolescents with CHD and healthy controls to determine how cortical metrics (CT, SA) drive the widely-reported reduction of brain cortical volume and cognitive impairments in CHD populations. We found total and regional reductions in both SA and CT in the CHD group, with SA affecting cortical volume reduction more than CT. Furthermore, across the whole sample, larger total and regional SA was strongly associated with higher IQ and EF. Mean CT was weakly associated with IQ but not with EF, and no significant associations were identified between regional CT and IQ/EF. In several regions, the association between cortical structures and IQ/EF were stronger in patients compared to controls.

### Reduction of CT and SA in patients with CHD

This is the first study to investigate whether a reduction in total and regional SA and CT contributes to lower cortical volumes in populations with CHD, and to report that total SA affected cortical volume more than mean CT, even in a sample including CHD, consistent with previous results in healthy adults.^33^ These findings provide more precise insights into how brain volume is affected in patients with CHD, as cortical volume is computed via the multiplication of SA and CT. Furthermore, we demonstrated that adolescents with CHD showed a significantly smaller total SA, and regionally in the left inferior parietal and postcentral regions, right middle temporal and rostral medial frontal and supramarginal regions, compared to controls. These detailed maps showing regional SA reductions are new, as previous morphometric studies in fetuses,^5^ neonates^16,29^ and adolescents^30^ all reported a reduction of total SA without regional specification and one adolescent study with lobe-based reduction of SA in temporal and parietal lobes, less so in frontal lobes, but not in occipital lobes.^20^

For the CT, we identified decreased mean CT in patients with CHD compared to controls globally and regionally, in the bilateral precuneus, precentral and rostral middle frontal, and the left inferior temporal region. Findings from previous studies on mean CT in populations with CHD are inconsistent. One study in neonates^16^ and another in adolescents^20^ found no statistically significant group difference in mean CT, while another adolescent/adult study found significantly decreased CT.^30^ Another study by Cordina examined regional CT in 10 adult patients (mean age: 40 years) and reported a bilateral decrease in the dorsolateral prefrontal cortex, the precentral gyrus, the superior, inferior and supramarginal gyri, precuneus, and the middle temporal gyrus.^31^ However, the regional CT reduction seen in our sample is less prominent than their results. This inconsistency in mean/regional CT results might stem from differences in sample size, age, CHD severity between the samples or from a lack of regional specificity, especially in mean CT studies.

Interestingly, we found that cyanotic CHD patients, compared to acyanotic CHD patients, and patients with univentricular defects, compared to those with biventricular defects, showed a statistically significantly lower mean CT, although the effect is relatively small. In contrast, no significant differences in SA or cortical volume were identified between CHD subtypes. Furthermore, the length of ICU stay after the first CPB surgery, which is an indirect indicator of CHD severity, was not associated with any of the global cortical metrics. This was unexpected, as a number of studies suggest CHD severity tends to be negatively associated with brain volume.^3,52^ Our CHD sample was quite heterogeneous, so the cortical structural differences among patients with CHD should be confirmed with a larger sample.

### Mechanism of reduction in CT and SA in patients with CHD

Different mechanisms may exist for reduced SA and CT seen in populations with CHD, as SA and CT are regulated by distinct genetic mechanisms,^24,25^ underscoring their independent developmental origins. SA is influenced by the number and spacing of cortical columns, determined by the proliferation of progenitor cells (symmetric division) in the subventricular zone (SVZ).^53–55^ In contrast, CT depends on the number of neurons within cortical columns, influenced by neurogenic (asymmetric) division.^53,54^ In addition to the number of neurons, CT is also influenced by the type and size of neurons, the dendritic and synaptic complexity, vascularisation and the amount of neuropil and non-neuronal elements such as microglia and astrocytes.^56–58^ Consequently, alterations of distinct early developmental processes may lead to different effects on SA and CT, which is evident in patients with CHD, who demonstrate a stronger reduction in SA than CT. CHD-related risk may specifically affect the progenitor cells in the SVZ, which is crucial for later expansion of the SA.

Indeed, animal studies support this hypothesis; researchers have been investigating the precise effects of CHD-related hypoperfusion and hypoxia on cortical development at the cellular level. Morton induced chronic hypoxia in perinatal piglets and found that hypoxia reduces proliferation and neurogenesis in the SVZ. This resulted in a depleted source of interneurons destined to populate the frontal cortex, which limited cortical expansion. This depletion of neuroblasts within SVZ was replicated with post-mortem neonates born with CHD.^59^ Another study investigating neonatal piglets also showed that cardiopulmonary bypass surgery-induced insult alone impaired postnatal neurogenesis and migration to the frontal lobe, leading to an imbalance of excitatory/inhibitory interneurons and restricted cortical expansion/maturation.^60^ Given that SA is mainly influenced by the proliferation of neural precursor cells and formation of cortical columns, and CT is influenced not only by neurogenesis, thus not by neuronal density alone, we can assume the impact of CHD might appear more consistent and evident for SA than CT due to this difference in ontological trajectories. However, among patients with CHD, more severe types (cyanotic, univentricular) showed lower mean CT, but not in total SA or cortical volume, compared to milder types. We could assume SA might be more genetically driven and reduced in all CHDs, while CT is more vulnerable to early hypoxia and altered blood perfusion (i.e., particularly reduced in the severe cases). Nonetheless, further investigations are needed as to why SA is more largely affected than CT in populations with CHD as a whole and why CT is affected especially among those with severe CHD types.

### Cognitive functions and cortical structures

#### SA and cognitive functions in healthy populations

We found that across the whole sample, larger total SA was associated with both higher IQ and EF. Regionally, SA was associated with IQ in the left medial orbitofrontal, isthmus cingulate and right superior temporal regions and with EF in the left postcentral and right pars opercularis regions. These findings are consistent with earlier studies reporting a positive correlation between SA and cognitive outcomes in healthy populations.^34,61–65^ For example, Waldhovd reported significant associations between general cognitive ability and SA in prefrontal, medial, and posterolateral temporal regions in children aged 4 to 12 years.^62^ Schmitt found positive correlations between SA expansion over time and IQ throughout bilateral hemispheres in youth, where SA-IQ associations were genetically mediated.^63^ Another study investigated children aged 9 to 11 years, reporting that greater SA in the left temporal and middle frontal regions, which are involved in language functions, was associated with higher crystallised intelligence (reading and vocabulary) scores. Conversely, larger SA in the anterior cingulate and insula cortices, implicated in cognitive control, was associated with higher fluid intelligence (novel problem-solving) scores.^34^ Colom also reported that larger SA in the right middle frontal gyrus was associated with fluid intelligence and working memory, while the inferior frontal gyrus was associated with crystallised intelligence in young adults.^65^ A similar study in adults aged 21 to 35 years showed that crystallised intelligence was linked to larger SA in the left inferior temporal and right middle temporal areas, whereas fluid intelligence was associated with regions including the left and right superior parietal and left caudal middle frontal areas.^64^ Despite the differences in cognitive measures and significant areas identified in previous studies, the association between total/regional SA and cognitive measures seems highly consistent, which aligns with our results.

#### CT and cognitive functions in healthy populations

In contrast to consistent associations between SA and cognitive functions, the relationships between mean/regional CT and cognitive functions were not straightforward in our study. Mean CT was positively associated with IQ but not with EF. Furthermore, no statistically significant regional CT clusters were associated with IQ or the EF. Our mixed results on mean and regional CT reflect the previous literature on this subject, where both significant and nonsignificant findings have been reported.

Several studies have reported little or no correlation between CT and cognitive outcomes. For example, Tamnes found no significant associations between mean CT and IQ in a sample aged 8–30 years.^66^ Similarly, mean or regional CT did not predict crystallised or fluid intelligence in children aged 9-11,^34^ young adults aged 21 to 35^64^ and young adults with a mean age of 19.9.^65^ On the other hand, some studies reported significant relationships between CT and cognition, although the direction and regional specificity of these associations vary widely. For instance, positive associations between CT and general intelligence have been observed in frontal, temporal, parietal, and occipital areas in samples aged 4-12,^62^ 6 to 18,^67^ 9 to 24,^68^ and children to adults with a mean age of 11.3.^69^ Choi found positive associations in the left temporal cortex and negative associations in the lateral parietal cortex between IQ and CT in young adults (mean 20.9 years).^70^ In contrast, only negative associations were found in a sample aged 9-11 between CT and general intelligence in the rostral cingulate and the right precuneus,^71^ and also in an adult sample aged 21 to 35, in the caudal middle frontal and superior parietal areas.^64^

This variability in findings on the CT-cognition relationship may arise from differences in MRI processing and cognitive measurement methods, but a more likely explanation is the dynamic nature of CT-cognition relationships across the lifespan. A longitudinal study by Shaw reported a primarily negative association between intelligence and CT in early childhood (3.8–8.4 years), which later shifted to a prominent positive one in late childhood (8.6–11.7 years).^72^ Sowell observed that greater vocabulary improvement from ages 5 to 11 was linked to cortical thinning in the frontal and parietal regions.^73^ Another study reported that CT and intelligence were not significantly associated at 9, but negative associations emerged by age 12.^74^ Schnack observed that in children aged 10 years, a higher IQ was associated with thinner cortices; however, this relationship reversed to a positive one by age 42.^61^ In contrast, Menary found that younger participants (9 to 16 years) demonstrated robust positive associations between CT and general intelligence, whereas older participants (16 to 24 years old) did not.^68^ Our cross-sectional sample aged 10-15 showed a positive correlation between mean CT and IQ, but no significant regional association, which may reflect a limited aspect of the age-dependent, evolving nature of CT-cognition relationships.

#### CHD-specific associations between cortical structures and cognitive functions

When investigating global structures, higher total SA, mean CT and cortical volume were associated with better IQ in both patients and controls, without a significant group interaction. However, when investigated regionally, the cortical associations with IQ/EF were stronger in patients with CHD than in controls in several areas. Those areas include: the left pars triangularis, lingual and the right lateral orbitofrontal regions (SA∼IQ), the left postcentral and the right pars opercularis regions (SA∼EF) the left rostral middle frontal region (CT∼IQ), the bilateral pars opercularis, the left lateral occipital regions (Vol∼IQ), the left superior temporal region (Vol∼EF).

Our findings are comparable to a recent study by Abboud, which investigated the associations between cortical features and EF assessed using a short behavioural rating questionnaire in youth with CHD (aged 16-32).^30^ They identified increased CT associated with worse EF in the precuneus, posterior language area, superior temporal, parietal, middle frontal, and anterior cingulate cortex in patients with CHD. These areas overlap with networks that are critical for EF, like cognitive flexibility and decision-making. They also observed reduced SA in the lateral sulcus, associated with a worse EF score.^30^ It is intriguing that they found increased CT linked with worse EF across the brain, while only one limited region of SA (lateral sulcus) was associated with EF. These results seem contradictory to our findings of strong SA associations with IQ/EF but not with CT. Another study identified a relationship between reduced CT and lower IQ in school-aged children with tetralogy of Fallot, in the left precuneus and right caudal middle frontal cortex.^32^ These previous findings and our results vary, possibly due to the dynamic change in CT-cognition relationship throughout the lifespan, as discussed previously. Given the consistent findings of positive SA-cognition relationships in previous studies of healthy populations, our sample with CHD is not an exception; thus, SA-cognition in CHD should be confirmed in future studies.

Furthermore, our findings on the stronger associations between cortical structures and cognitive function in patients compared to controls align with previous CHD studies, specifically regarding total/regional brain volumes.^20,75^ This suggests that although there are strong associations between cortical structures and cognitive function in healthy populations, the associations are even more apparent in populations with CHD, when brain growth is limited by a disease process.

#### Is SA a better indicator than CT for cognitive function?

Our results suggest that SA might be a better indicator for cognitive functions than CT, as correlations with IQ/EF were widely seen only in SA, but not in CT. Theoretically, higher cortical SA can enhance information processing capacity as larger SA equals a higher number of cortical columns, which are the functional units of the cortex.^22,76^ These additional columns allow for greater functional specificity of cortical columns and reduced overlap in neural representations, enabling the brain to process and store information more efficiently.^77^ This increased SA for distinct neural representations is also shown in the early visual cortex (V1 and V2), where visual SA correlated positively with neural population tuning sharpness and perceptual discrimination accuracy, while increased CT has the opposite effect.^78^ Furthermore, while CT contributes to cognitive function by reflecting interlaminar connections and synaptic remodelling, it might fluctuate depending on experience and learning,^79^ or show thinning during childhood due to the reflection of myelination.^80^ With previous literature on SA-cognition relationships and ontogeny, larger cortical SA seems to be the consistent underlying factor for better cognitive performance rather than higher CT.^64^

### Limitations

There are several limitations to this study. Although our sample size was sufficient to detect CHD-control group differences in cortical metrics, the limited sample size and heterogeneity of CHD diagnoses affected the generalisability of this study. We created a custom template for our adolescents for better anatomical representation, more accurate registration, and potentially more robust statistical analyses. However, the use of the custom template might have made our results study-specific and slightly less comparable to other studies with the standard fsaverage template. Furthermore, elucidating the longitudinal relationship between cortical structure and cognitive functions was beyond the scope of our cross-sectional study, although earlier studies on healthy populations indicated lifespan changes in the structure-function relationship.^61,72^ In future studies, our findings on the cortical structural association with cognitive outcomes in adolescents with CHD should be confirmed with a larger sample size, and changes in the associations over time should be assessed in longitudinal studies.

## Data availability

The de-identified data are available from the corresponding author upon reasonable request.

## Acknowledgements

We would like to thank all the children, adolescents and adults who participated in this study. We would like to thank Nadja Naef and Alenka Schmid for their valuable support in performing the MRI scans.

## Reference

1. Bernier PL, Stefanescu A, Samoukovic G, Tchervenkov CI. The challenge of congenital heart disease worldwide: epidemiologic and demographic facts. Semin Thorac Cardiovasc Surg Pediatr Card Surg Annu. 2010;13(1):26–34. doi:10.1053/j.pcsu.2010.02.005

2. Aleksonis HA, King TZ. Relationships Among Structural Neuroimaging and Neurocognitive Outcomes in Adolescents and Young Adults with Congenital Heart Disease: A Systematic Review. Neuropsychol Rev. Published online July 1, 2022. doi:10.1007/s11065-022-09547-2

3. Brossard-Racine M, Panigrahy A. Structural Brain Alterations and Their Associations With Function in Children, Adolescents, and Young Adults With Congenital Heart Disease. Can J Cardiol. 2023;39(2):123–132. doi:10.1016/j.cjca.2022.10.028

4. Sun L, Macgowan CK, Sled JG, et al. Reduced fetal cerebral oxygen consumption is associated with smaller brain size in fetuses with congenital heart disease. Circulation. 2015;131(15):1313–1323. doi:10.1161/CIRCULATIONAHA.114.013051

5. Clouchoux C, du Plessis AJ, Bouyssi-Kobar M, et al. Delayed cortical development in fetuses with complex congenital heart disease. Cereb Cortex. 2013;23(12):2932–2943. doi:10.1093/cercor/bhs281

6. Limperopoulos C, Tworetzky W, McElhinney DB, et al. Brain volume and metabolism in fetuses with congenital heart disease: evaluation with quantitative magnetic resonance imaging and spectroscopy: Evaluation with quantitative magnetic resonance imaging and spectroscopy. Circulation. 2010;121(1):26–33. doi:10.1161/CIRCULATIONAHA.109.865568

7. Rollins CK, Ortinau CM, Stopp C, et al. Regional Brain Growth Trajectories in Fetuses with Congenital Heart Disease. Ann Neurol. 2021;89(1):143–157. doi:10.1002/ana.25940

8. Masoller N, Sanz-Cortés M, Crispi F, et al. Severity of fetal brain abnormalities in congenital heart disease in relation to the main expected pattern of in utero brain blood supply. Fetal Diagn Ther. 2016;39(4):269–278. doi:10.1159/000439527

9. Schellen C, Ernst S, Gruber GM, et al. Fetal MRI detects early alterations of brain development in Tetralogy of Fallot. Am J Obstet Gynecol. 2015;213(3):392.e1-7. doi:10.1016/j.ajog.2015.05.046

10. Lee VK, Ceschin R, Reynolds WT, et al. Postnatal brain trajectories and maternal Intelligence predict childhood outcomes in complex CHD. J Clin Med. 2024;13(10):2922. doi:10.3390/jcm13102922

11. Ortinau CM, Mangin-Heimos K, Moen J, et al. Prenatal to postnatal trajectory of brain growth in complex congenital heart disease. NeuroImage Clin. 2018;20:913–922. doi:10.1016/j.nicl.2018.09.029

12. Neukomm A, Ehrler M, Feldmann M, et al. Perioperative course and socioeconomic status predict long-term neurodevelopment better than perioperative conventional neuroimaging in children with congenital heart disease. J Pediatr. Published online August 7, 2022. doi:10.1016/j.jpeds.2022.07.032

13. Meuwly E, Feldmann M, Knirsch W, et al. Postoperative brain volumes are associated with one-year neurodevelopmental outcome in children with severe congenital heart disease. Sci Rep. 2019;9(1):10885. doi:10.1038/s41598-019-47328-9

14. Cromb D, Bonthrone AF, Maggioni A, et al. Individual Assessment of Perioperative Brain Growth Trajectories in Infants With Congenital Heart Disease: Correlation With Clinical and Surgical Risk Factors. J Am Heart Assoc. 2023;12(14):e028565. doi:10.1161/JAHA.122.028565

15. Heye KN, Knirsch W, Latal B, et al. Reduction of brain volumes after neonatal cardiopulmonary bypass surgery in single-ventricle congenital heart disease before Fontan completion. Pediatr Res. 2018;83(1-1):63–70. doi:10.1038/pr.2017.203

16. Claessens NHP, Moeskops P, Buchmann A, et al. Delayed cortical gray matter development in neonates with severe congenital heart disease. Pediatr Res. 2016;80(5):668–674. doi:10.1038/pr.2016.145

17. von Rhein M, Buchmann A, Hagmann C, et al. Severe congenital heart defects are associated with global reduction of neonatal brain volumes. J Pediatr. 2015;167(6):1259–63.e1. doi:10.1016/j.jpeds.2015.07.006

18. Naef N, Schlosser L, Brugger P, et al. Brain volumes in adults with congenital heart disease correlate with executive function abilities. Brain Imaging Behav. 2021;15(5):2308–2316. doi:10.1007/s11682-020-00424-1

19. Verrall CE, Yang JYM, Chen J, et al. Neurocognitive dysfunction and smaller brain volumes in adolescents and adults with a Fontan circulation. Circulation. 2021;143(9):878–891. doi:10.1161/CIRCULATIONAHA.120.048202

20. von Rhein M, Buchmann A, Hagmann C, et al. Brain volumes predict neurodevelopment in adolescents after surgery for congenital heart disease. Brain. 2014;137(Pt 1):268–276. doi:10.1093/brain/awt322

21. Hiraiwa A, Kawasaki Y, Ibuki K, et al. Brain Development of Children With Single Ventricle Physiology or Transposition of the Great Arteries: A Longitudinal Observation Study. Semin Thorac Cardiovasc Surg. 2020;32(4):936–944. doi:10.1053/j.semtcvs.2019.06.013

22. Mountcastle VB. The columnar organization of the neocortex. Brain. 1997;120 (Pt 4)(4):701–722. doi:10.1093/brain/120.4.701

23. Roth G, Dicke U. Evolution of the brain and intelligence. Trends Cogn Sci. 2005;9(5):250–257. doi:10.1016/j.tics.2005.03.005

24. Panizzon MS, Fennema-Notestine C, Eyler LT, et al. Distinct genetic influences on cortical surface area and cortical thickness. Cereb Cortex. 2009;19(11):2728–2735. doi:10.1093/cercor/bhp026

25. Grasby KL, Jahanshad N, Painter JN, et al. The genetic architecture of the human cerebral cortex. Science. 2020;367(6484). doi:10.1126/science.aay6690

26. Mechelli A, Price C, Friston K, Ashburner J. Voxel-based morphometry of the human brain: Methods and applications. Curr Med Imaging Rev. 2005;1(2):105–113. doi:10.2174/1573405054038726

27. Hutton C, Draganski B, Ashburner J, Weiskopf N. A comparison between voxel-based cortical thickness and voxel-based morphometry in normal aging. Neuroimage. 2009;48(2):371–380. doi:10.1016/j.neuroimage.2009.06.043

28. Ortinau CM, Rollins CK, Gholipour A, et al. Early-Emerging Sulcal Patterns Are Atypical in Fetuses with Congenital Heart Disease. Cereb Cortex. 2019;29(8):3605–3616. doi:10.1093/cercor/bhy235

29. Ortinau C, Alexopoulos D, Dierker D, Van Essen D, Beca J, Inder T. Cortical folding is altered before surgery in infants with congenital heart disease. J Pediatr. 2013;163(5):1507–1510. doi:10.1016/j.jpeds.2013.06.045

30. Abboud F, Easson K, Ehrler M, et al. Cortical alterations associated with executive function deficits in youth with a congenital heart defect. Imaging Neuroscience. 2024;2:1–17. doi:10.1162/imag_a_00371

31. Cordina R, Grieve S, Barnett M, Lagopoulos J, Malitz N, Celermajer DS. Brain volumetric, regional cortical thickness and radiographic findings in adults with cyanotic congenital heart disease. Neuroimage Clin. 2014;4:319–325. doi:10.1016/j.nicl.2013.12.011

32. Ma S, Li Y, Liu Y, et al. Changes in cortical thickness are associated with cognitive ability in postoperative school-aged children with tetralogy of Fallot. Front Neurol. 2020;11:691. doi:10.3389/fneur.2020.00691

33. Im K, Lee JM, Lyttelton O, Kim SH, Evans AC, Kim SI. Brain size and cortical structure in the adult human brain. Cereb Cortex. 2008;18(9):2181–2191. doi:10.1093/cercor/bhm244

34. Palmer CE, Zhao W, Loughnan R, et al. Distinct regionalization patterns of cortical morphology are associated with cognitive performance across different domains. Cereb Cortex. 2021;31(8):3856–3871. doi:10.1093/cercor/bhab054

35. Ehrler M, Naef N, Tuura RO, Latal B. Executive function and brain development in adolescents with severe congenital heart disease (Teen Heart Study): protocol of a prospective cohort study. BMJ Open. 2019;9(10):e032363. doi:10.1136/bmjopen-2019-032363

36. Largo RH, Pfister D, Molinari L, Kundu S, Lipp A, Duc G. Significance of prenatal, perinatal and postnatal factors in the development of AGA preterm infants at five to seven years. Dev Med Child Neurol. 1989;31(4):440–456. doi:10.1111/j.1469-8749.1989.tb04022.x

37. Waldmann HC. Kurzformen des HAWIK-IV: Statistische Bewertung in verschiedenen Anwendungsszenarien. Diagnostica. 2008;54(4):202–210. doi:10.1026/0012-1924.54.4.202

38. Ehrler M, Latal B, Polentarutti S, von Rhein M, Held L, Wehrle FM. Pitfalls of using IQ short forms in neurodevelopmental disorders: a study in patients with congenital heart disease. Pediatr Res. 2020;87(5):917–923. doi:10.1038/s41390-019-0667-2

39. Delis DC, Kaplan E, Kramer JH. Delis-Kaplan Executive Function System. Assessment. doi:10.1037/t15082-000

40. Zimmermann P, Fimm B. Testbatterie zur Erfassung von Aufmerksamkeitsstörungen (TAP 2.3.). Published online 2012.

41. Aschenbrenner S, Tucha O, Lange KW. Regensburger Wortflüssigkeits-Test: RWT. Hogrefe, Verlag für Psychologie; 2000.

42. von Werdt L, Binz TM, O’Gorman RT, et al. Stress Markers, Executive Functioning, and Resilience Among Early Adolescents With Complex Congenital Heart Disease. JAMA Netw Open. 2024;7(2):e2355373. doi:10.1001/jamanetworkopen.2023.55373

43. Fischl B, Sereno MI, Dale AM. Cortical surface-based analysis. II: Inflation, flattening, and a surface-based coordinate system. Neuroimage. 1999;9(2):195–207. doi:10.1006/nimg.1998.0396

44. Dale AM, Fischl B, Sereno MI. Cortical surface-based analysis. I. Segmentation and surface reconstruction. Neuroimage. 1999;9(2):179-194. doi:10.1006/nimg.1998.0395

45. Fischl B, Sereno MI, Tootell RB, Dale AM. High-resolution intersubject averaging and a coordinate system for the cortical surface. Hum Brain Mapp. 1999;8(4):272–284. doi:10.1002/(sici)1097-0193(1999)8:4<272::aid-hbm10>3.0.co;2-4

46. Desikan RS, Ségonne F, Fischl B, et al. An automated labeling system for subdividing the human cerebral cortex on MRI scans into gyral based regions of interest. Neuroimage. 2006;31(3):968–980. doi:10.1016/j.neuroimage.2006.01.021

47. Han X, Jovicich J, Salat D, et al. Reliability of MRI-derived measurements of human cerebral cortical thickness: the effects of field strength, scanner upgrade and manufacturer. Neuroimage. 2006;32(1):180–194. doi:10.1016/j.neuroimage.2006.02.051

48. Jovicich J, Czanner S, Han X, et al. MRI-derived measurements of human subcortical, ventricular and intracranial brain volumes: Reliability effects of scan sessions, acquisition sequences, data analyses, scanner upgrade, scanner vendors and field strengths. Neuroimage. 2009;46(1):177–192. doi:10.1016/j.neuroimage.2009.02.010

49. R core team. R: A Language and Environment for Statistical Computing.; 2023. https://www.R-project.org/

50. Huisenga D, La Bastide-Van Gemert S, Van Bergen A, Sweeney J, Hadders-Algra M. Developmental outcomes after early surgery for complex congenital heart disease: a systematic review and meta-analysis. Dev Med Child Neurol. 2021;63(1):29–46. doi:10.1111/dmcn.14512

51. Greve DN, Fischl B. False positive rates in surface-based anatomical analysis. Neuroimage. 2018;171:6–14. doi:10.1016/j.neuroimage.2017.12.072

52. Bonthrone AF, Kelly CJ, Ng IHX, Counsell SJ. MRI studies of brain size and growth in individuals with congenital heart disease. Transl Pediatr. 2021;10(8):2171–2181. doi:10.21037/tp-20-282

53. Rakic P. Specification of cerebral cortical areas. Science. 1988;241(4862):170–176. doi:10.1126/science.3291116

54. Rakic P. A small step for the cell, a giant leap for mankind: a hypothesis of neocortical expansion during evolution. Trends Neurosci. 1995;18(9):383–388. doi:10.1016/0166-2236(95)93934-p

55. Chenn A, Walsh CA. Increased neuronal production, enlarged forebrains and cytoarchitectural distortions in beta-catenin overexpressing transgenic mice. Cereb Cortex. 2003;13(6):599–606. doi:10.1093/cercor/13.6.599

56. Vidal-Pineiro D, Parker N, Shin J, et al. Cellular correlates of cortical thinning throughout the lifespan. Sci Rep. 2020;10(1):21803. doi:10.1038/s41598-020-78471-3

57. Carlo CN, Stevens CF. Structural uniformity of neocortex, revisited. Proc Natl Acad Sci U S A. 2013;110(4):1488–1493. doi:10.1073/pnas.1221398110

58. Huttenlocher PR, Dabholkar AS. Regional differences in synaptogenesis in human cerebral cortex. J Comp Neurol. 1997;387(2):167–178. doi:10.1002/(sici)1096-9861(19971020)387:2<167::aid-cne1>3.0.co;2-z

59. Morton PD, Korotcova L, Lewis BK, et al. Abnormal neurogenesis and cortical growth in congenital heart disease. Sci Transl Med. 2017;9(374). doi:10.1126/scitranslmed.aah7029

60. Dhari Z, Leonetti C, Lin S, et al. Impact of cardiopulmonary bypass on neurogenesis and cortical maturation. Ann Neurol. 2021;90(6):913–926. doi:10.1002/ana.26235

61. Schnack HG, van Haren NEM, Brouwer RM, et al. Changes in thickness and surface area of the human cortex and their relationship with intelligence. Cereb Cortex. 2015;25(6):1608–1617. doi:10.1093/cercor/bht357

62. Walhovd KB, Krogsrud SK, Amlien IK, et al. Neurodevelopmental origins of lifespan changes in brain and cognition. Proc Natl Acad Sci U S A. 2016;113(33):9357–9362. doi:10.1073/pnas.1524259113

63. Schmitt JE, Neale MC, Clasen LS, et al. A comprehensive quantitative genetic analysis of cerebral surface area in youth. J Neurosci. 2019;39(16):3028–3040. doi:10.1523/JNEUROSCI.2248-18.2019

64. Tadayon E, Pascual-Leone A, Santarnecchi E. Differential Contribution of Cortical Thickness, Surface Area, and Gyrification to Fluid and Crystallized Intelligence. Cereb Cortex. 2020;30(1):215–225. doi:10.1093/cercor/bhz082

65. Colom R, Burgaleta M, Román FJ, et al. Neuroanatomic overlap between intelligence and cognitive factors: morphometry methods provide support for the key role of the frontal lobes. Neuroimage. 2013;72:143–152. doi:10.1016/j.neuroimage.2013.01.032

66. Tamnes CK, Fjell AM, Østby Y, et al. The brain dynamics of intellectual development: waxing and waning white and gray matter. Neuropsychologia. 2011;49(13):3605–3611. doi:10.1016/j.neuropsychologia.2011.09.012

67. Karama S, Ad-Dab’bagh Y, Haier RJ, et al. Positive association between cognitive ability and cortical thickness in a representative US sample of healthy 6 to 18 year-olds. Intelligence. 2009;37(2):145–155. doi:10.1016/j.intell.2008.09.006

68. Menary K, Collins PF, Porter JN, et al. Associations between cortical thickness and general intelligence in children, adolescents and young adults. Intelligence. 2013;41(5):597–606. doi:10.1016/j.intell.2013.07.010

69. Schmitt JE, Raznahan A, Clasen LS, et al. The dynamic associations between cortical thickness and general intelligence are genetically mediated. Cereb Cortex. 2019;29(11):4743–4752. doi:10.1093/cercor/bhz007

70. Choi YY, Shamosh NA, Cho SH, et al. Multiple bases of human intelligence revealed by cortical thickness and neural activation. J Neurosci. 2008;28(41):10323–10329. doi:10.1523/JNEUROSCI.3259-08.2008

71. Zhao Q, Voon V, Zhang L, Shen C, Zhang J, Feng J. The ABCD study: Brain heterogeneity in intelligence during a neurodevelopmental transition stage. Cereb Cortex. 2022;32(14):3098–3109. doi:10.1093/cercor/bhab403

72. Shaw P, Greenstein D, Lerch J, et al. Intellectual ability and cortical development in children and adolescents. Nature. 2006;440(7084):676-679. doi:10.1038/nature04513

73. Sowell ER, Thompson PM, Leonard CM, Welcome SE, Kan E, Toga AW. Longitudinal mapping of cortical thickness and brain growth in normal children. J Neurosci. 2004;24(38):8223–8231. doi:10.1523/JNEUROSCI.1798-04.2004

74. Brouwer RM, van Soelen ILC, Swagerman SC, et al. Genetic associations between intelligence and cortical thickness emerge at the start of puberty. Hum Brain Mapp. 2014;35(8):3760–3773. doi:10.1002/hbm.22435

75. Latal B, Patel P, Liamlahi R, Knirsch W, O’Gorman Tuura R, von Rhein M. Hippocampal volume reduction is associated with intellectual functions in adolescents with congenital heart disease. Pediatr Res. 2016;80(4):531–537. doi:10.1038/pr.2016.122

76. Rakic P. Evolution of the neocortex: a perspective from developmental biology. Nat Rev Neurosci. 2009;10(10):724–735. doi:10.1038/nrn2719

77. Ringo JL. Neuronal interconnection as a function of brain size. Brain Behav Evol. 1991;38(1):1–6. doi:10.1159/000114375

78. Song C, Schwarzkopf DS, Kanai R, Rees G. Neural population tuning links visual cortical anatomy to human visual perception. Neuron. 2015;85(3):641–656. doi:10.1016/j.neuron.2014.12.041

79. Wenger E, Brozzoli C, Lindenberger U, Lövdén M. Expansion and renormalization of human brain structure during skill acquisition. Trends Cogn Sci. 2017;21(12):930–939. doi:10.1016/j.tics.2017.09.008

80. Natu VS, Gomez J, Barnett M, et al. Apparent thinning of human visual cortex during childhood is associated with myelination. Proc Natl Acad Sci U S A. 2019;116(41):20750–20759. doi:10.1073/pnas.1904931116

