## Supplemental material for "Cortical surface area drives volumetric and cognitive deficits in complex congenital heart disease"

**Supplemental Table 1**
**Neuropsychological Test Battery to Assess Executive Functions Performance**

| Domains | Neuropsychological test | Test measurement |
|---|---|---|
| Working memory | Digit span forward and backward (WISC-IV) | No. of correct items |
| | Letter-number sequencing (WISC-IV) | No. of correct items |
| | Corsi block tapping test (Corsi) | No. of correct items |
| Inhibition | Inhibition Subtest interference, colour word interference task (D-KEFS) | Completion time |
| | Go/NoGo (TAP) | No. of commission errors |
| Cognitive flexibility | Subtest letter-number-switching, trail making task(D-KEFS) | Completion time |
| | TAP flexibility (TAP) | Median reaction time |
| Fluency | Subtests s-words and animals (RWT) | No. of correct items |
| | Subtest filled-dots-only, design fluency test (D-KEFS) | No. of correct items |
| Planning | Tower task (D-KEFS) | Total achievement score |

**Supplemental Table 2**
**Diagnosis of congenital heart disease**

| Diagnosis | number |
|---|---|
| Aortic stenosis valvular | 1 |
| Atrioventricular septal defect (AVSD) | 1 |
| Complete AVSD | 2 |
| Coarctation of the aorta (CoA) | 2 |
| d-Transposition of the great arteries (d-TGA) | 16 |
| Double inlet left ventricle (DILV) | 1 |
| Ebstein's anomaly (severe) + Pulmonary stenosis (PS) | 1 |
| Hypoplastic left heart syndrome (HLHS) | 4 |
| Pulmonary atresia (PA) | 1 |
| PA with intact ventricular septum | 1 |
| PA with VSD and aortopulmonary collaterals (MAPCAs) | 1 |
| Pulmonary stenosis valvular | 1 |
| Tricuspid atresia (TA) | 1 |
| Truncus arteriosus type 1 | 2 |
| Total anomalous pulmonary venous connection (TAPVC) | 2 |
| Tetralogy of Fallot (TOF) | 4 |
| ventricular septal defect (VSD) doubly committed | 1 |
| Perimembranous VSD | 6 |
| Subaortic VSD | 1 |

**Supplemental Table 3**
**Group difference (CHD vs. controls) in cortical metrics**

| Effect | B | Standard error | 95% CI | β | p-value |
|---|---|---|---|---|---|
| **Cortical volume** | | | | | |
| group | 34.833 | 9.483 | 0.136 ~ 0.430 | 0.283 | 5.91E-04 |
| age | -5.950 | 3.204 | -0.270 ~ 0.006 | -0.132 | 0.082 |
| sex | -56.627 | 8.223 | -0.597 ~ -0.347 | -0.472 | 4.23E-10 |
| SES | 7.000 | 1.993 | 0.119 ~ 0.406 | 0.262 | 0.001 |
| **lh mean CT** | | | | | |
| group | 0.058 | 0.016 | 0.154 ~ 0.490 | 0.322 | 6.53E-04 |
| age | -0.020 | 0.005 | -0.463 ~ -0.154 | -0.309 | 4.14E-04 |
| sex | -0.025 | 0.014 | -0.294 ~ 0.013 | -0.141 | 0.079 |
| SES | -0.004 | 0.003 | -0.280 ~ 0.056 | -0.112 | 0.221 |
| **rh mean CT** | | | | | |
| group | 0.048 | 0.016 | 0.095 ~ 0.427 | 0.261 | 0.004 |
| age | -0.025 | 0.005 | -0.521 ~ -0.225 | -0.373 | 1.62E-05 |
| sex | -0.029 | 0.014 | -0.311 ~ -0.011 | -0.161 | 0.045 |
| SES | -0.001 | 0.003 | -0.198 ~ 0.131 | -0.034 | 0.692 |
| **lh total SA** | | | | | |
| group | 50.480 | 18.961 | 0.058 ~ 0.363 | 0.210 | 0.010 |
| age | -7.460 | 6.406 | -0.227 ~ 0.057 | -0.085 | 0.274 |
| sex | -112.064 | 16.442 | -0.606 ~ -0.353 | -0.479 | 4.54E-10 |
| SES | 16.081 | 3.986 | 0.164 ~ 0.455 | 0.310 | 1.58E-04 |
| **rh total SA** | | | | | |
| group | 48.678 | 18.929 | 0.052 ~ 0.359 | 0.205 | 0.011 |
| age | -5.964 | 6.395 | -0.212 ~ 0.074 | -0.069 | 0.353 |
| sex | -111.975 | 16.414 | -0.612 ~ -0.358 | -0.485 | 4.54E-10 |
| SES | 15.323 | 3.979 | 0.151 ~ 0.446 | 0.299 | 2.68E-04 |

Note: CT = cortical thickness (mm), SA = surface area ($cm^2$), lh = left hemisphere, rh= right hemisphere, CI = confidence interval, p-values are FDR corrected.

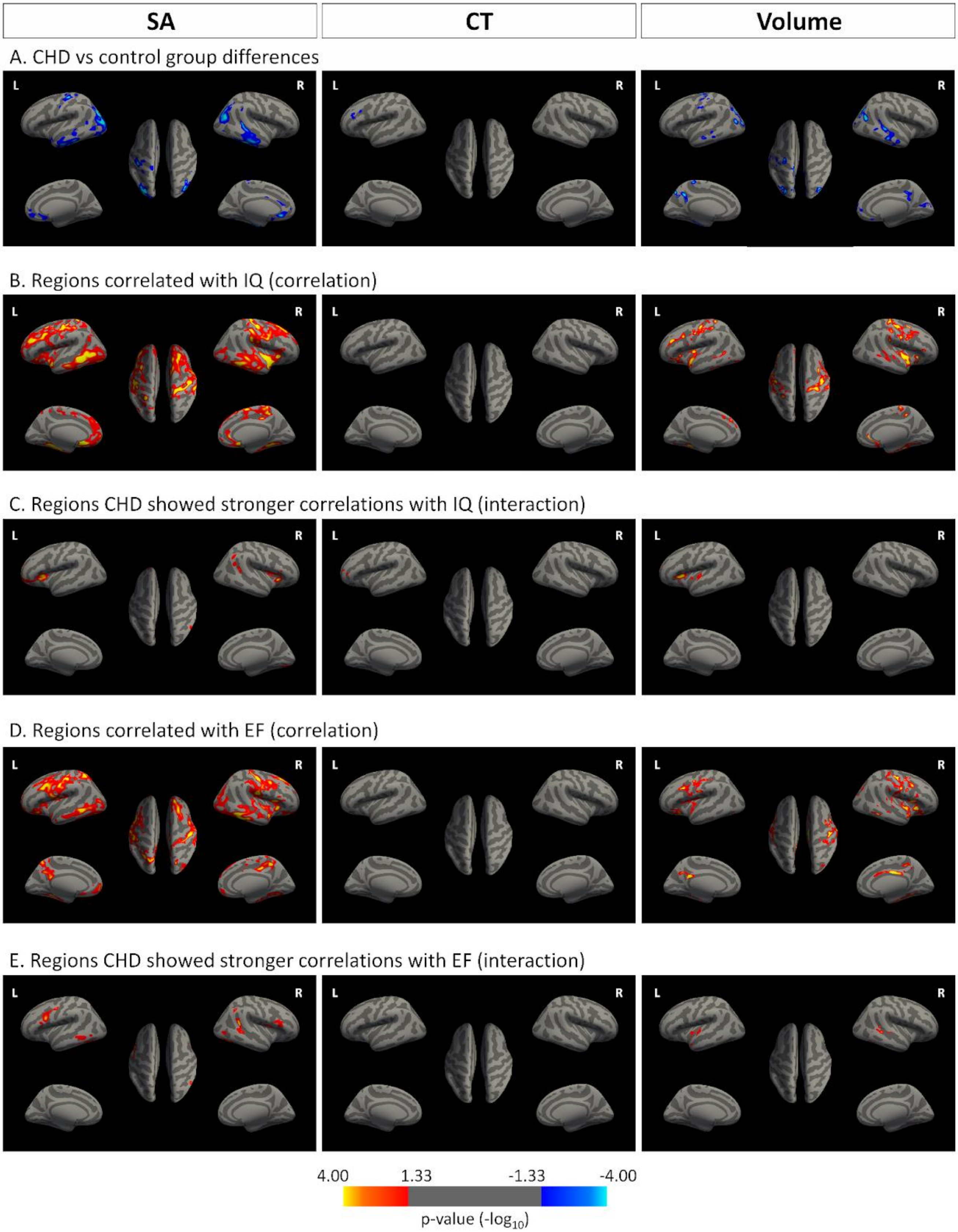

SA
CT
Volume
A. CHD vs control group differences
B. Regions correlated with IQ (correlation)
C. Regions CHD showed stronger correlations with IQ (interaction)
D. Regions correlated with EF (correlation)
E. Regions CHD showed stronger correlations with EF (interaction)
L
R
4.00
1.33
-1.33
-4.00
p-value ($-\log_{10}$)

**Supplemental Figure 1**
**Cortical surface structures with restricted subsample without any minor movement artefacts (N=111)**

This selected sample did not differ significantly compared to the main result (N=129), but the group differences turned out less widespread, especially for the CT. All analyses were adjusted for age and sex, $p < 0.05$, cluster-wise corrected, represented by blue clusters (negative correlation) or yellow-red clusters (positive correlation). The colour bar represents uncorrected significance values masked by the clusters that survived correction for multiple comparisons. SA = surface area, CT = cortical thickness, Volume = cortical volume. EF = executive function
**A.** Patients with CHD showed significantly lower SA than controls in the bilateral inferior temporal and medial orbitofrontal, left postcentral, the right inferior parietal regions (the leftmost), significantly lower CT in the left rostral middle frontal region (the middle figure), and significantly lower volume in bilateral middle temporal and precuneus, the left superior parietal, precentral and inferior temporal, the right inferior parietal, cuneus, lateral orbitofrantal, superior frontal regions (the rightmost). No significant positive clusters (yellow-red) were found.
**B.** SA in the left medial orbitofrontal, the right superior temporal and rostal anterior cingulate regions were associated with IQ (the leftmost). No significant clusters associated with IQ were found for CT (the middle). For cortical volume, the bilateral fusiform, superior temporal, the left rostral middle frontal, lateral occipital, middle temporal, superior frontal region and the right superior temporal regions were associated with IQ (the rightmost). No significant negative clusters (blue) were found.
**C.** The CHD group showed stronger associations between IQ and SA in the bilateral lateral orbitofrontal region and the lingual, and inferior parietal regions (the leftmost). For CT, the left frontal pole showed a stronger correlation with IQ in the CHD than the control group (middle). For volume, the left pars triangularis and rostral middle frontal regions showed stronger associations with IQ (the rightmost)
**D.** SA in the bilateral postcentral region was associated with the EF summary score (the leftmost). No significant clusters associated with EF summary score were found for CT (the middle). For cortical volume, the left caudal middle frontal,lateral orbitofrontal, fusiform, isthmus cingulate, the right pars opercularis, parahippocampal, posterior cingulate, inferior temporal, inferior parietal regions were associated with the EF summary score (the rightmost). No significant negative clusters (blue) were found.
**E.** The CHD group showed stronger associations between EF summary score and SA in the left middle temporal, pars opercularis, the right inferior parietal and precentral regions (the leftmost). For CT, there were no clusters where the group difference in EF summary score was significant (middle). For volume, the left superior temporal and right bankssts and fusiform region showed stronger associations with EF summary score than controls (the rightmost)
